## Supplement for "Laboratory diagnostic of acute kidney injury and its progression: risk of underdiagnosis in female and elderly patients"

**Text S1. Recalculation of Creatinine**

Estimated glomerular filtration rate (eGFR, ml/min/1.73m²) values at ULMC are calculated on the basis of the CKI-EPI equations [17] (using SCr in µmol/L) for a predominantly non-black cohort thus:

| Sex-specific knot mg/dL (µmol/L) | Equation |
|---|---|
| Females ≤ 0.7 (62) | $eGFR = 144 \times (SCr/0.7)^{-0.329} \times (0.993)^{Age}$ |
| Females > 0.7 (62) | $eGFR = 144 \times (SCr/0.7)^{-1.209} \times (0.993)^{Age}$ |
| Males ≤ 0.9 (80) | $eGFR = 141 \times (SCr/0.9)^{-0.411} \times (0.993)^{Age}$ |
| Males > 0.9 (80) | $eGFR = 141 \times (SCr/0.9)^{-1.209} \times (0.993)^{Age}$ |

For the reverse action, the inference of serum creatinine (SCr) from eGFR, we rearranged the equations to the following while retaining the original grouping below and at/above the sex-specific knot, respectively. The factors 0.9 (instead of 0.7), 141 (instead of 144), and -0.411 (instead of -0.329) are needed to transform a “female” SCr into “male” SCr via the original eGFR. The recalculation of males into younger individuals was achieved by setting the variable Age to 30.

| | |
|---|---|
| Below/at sex-specific knot | SCr = e ^ (ln(eGFR/141/0.993 ^ Age) / (-0.411)) * 0.9 * 88.42 |
| above sex-specific knot | SCr = e ^ (ln(eGFR/141/0.993 ^ Age) / (-1.209)) * 0.9 * 88.42 |

**Table S1. Comparison of progressive and non-progressive cases at first AKI detection during hospitalization in females and males. Variables are given as medians [interquartile range] or percentages.**

| | Female | | | Male | | |
|---|---|---|---|---|---|---|
| | **Progressive AKI** | **Non-progressive AKI** | **p-value** | **Progressive AKI** | **Non-progressive AKI** | **p-value** |
| Incidence proportion, n, % | 554, 22.0 | 2514, 78.0 | | 904, 25.0 | 3620, 75.0 | |
| **Basic patient characteristics** | | | | | | |
| Age (years) | 70.7 [59.0–79.5] | 72.4 [60.7–81.1] | **0.013** | 66.0 [57.0–75.9] | 68.0 [58.2–78.1] | **0.003** |
| Total length of hospitalization (days) | 24.7 [14.2–43.1] | 15.1 [8.3–27.1] | **< 0.001** | 26.3 [14.9–42.6] | 16.0 [8.2–28.3] | **< 0.001** |
| First eGFR | 58.0 [36.1–85.1] | 60.2 [36.0–85.5] | 0.696 | 61.9 [37.7–88.1] | 63.7 [39.7–88.2] | 0.371 |
| Last eGFR | 44.9 [25.2–74.4] | 54.9 [34.4–83.6] | **< 0.001** | 58.1 [38.1–85.7] | 46.0 [25.7–75.7] | **< 0.001** |
| Time to first AKI during hospitalization (days) | 4.9 [1.9–11.9] | 4.6 [1.9–10.6] | 0.242 | 4.5 [1.8–9.4] | 4.5 [1.9–10.0] | 0.336 |
| In-hospital mortality | 45.5 | 16.8 | **< 0.001** | 45.0 | 17.5 | **< 0.001** |
| **Comorbidities** | | | | | | |
| I10.- Hypertension | 47.7 | 48.3 | 0.823 | 46.9 | 46.2 | 0.750 |
| E11.- Diabetes mellitus | 31.4 | 31.9 | 0.847 | 35.2 | 34.2 | 0.607 |
| E86.- Exsiccosis | 6.0 | 4.5 | 0.176 | 3.4 | 4.6 | 0.143 |
| R57.- Shock | 37.9 | 12.6 | **< 0.001** | 44.3 | 16.8 | **< 0.001** |
| I25.- Coronary heart disease | 11.2 | 14.0 | 0.097 | 23.5 | 23.3 | 0.980 |
| I21.- Myocardial infarction | 3.4 | 3.2 | 0.869 | 6.2 | 4.5 | **0.046** |
| I50.- Cardiac insufficiency | 33.9 | 27.8 | **0.004** | 34.6 | 26.4 | **< 0.001** |
| A41.- Sepsis | 38.8 | 15.6 | **< 0.001** | 41.9 | 20.7 | **< 0.001** |
| K74.- Liver cirrhosis | 8.7 | 4.3 | **< 0.001** | 9.0 | 4.1 | **< 0.001** |

**Table S2. Comparison of age groups with regard to AKI stages and sex. Based on common AKI cases for females and females as male.**

| Age group | | First AKI stage during hospitalization, %, (n) | | | Maximum AKI stage during hospitalization, %, (n) | | |
|---|---|---|---|---|---|---|---|
| | | AKIN1 | AKIN2 | AKIN3 | AKIN1 | AKIN2 | AKIN3 |
| [18–41) | Female | 78.2 (140) | 15.1 (27) | **6.7 (12)** | 62.6 (112) | 20.1 (36) | **17.3 (31)** |
| | Female as male | 74.3 (133) | 14.0 (25) | **11.7 (21)** | 59.8 (107) | 13.4 (24) | **26.8 (48)** |
| [41–61) | Female | 81.3 (443) | 12.1 (66) | **6.6 (36)** | 61.3 (334) | 22.6 (123) | **16.1 (88)** |
| | Female as male | 83.1 (453) | 10.8 (59) | **6.1 (33)** | 62.2 (339) | 20.7 (113) | **17.1 (93)** |
| [61–81) | Female | 82.8 (1207) | 11.5 (167) | **5.7 (83)** | 65.5 (955) | 20.9 (305) | **13.5 (197)** |
| | Female as male | 83.1 (1211) | 10.3 (150) | **6.7 (97)** | 65.4 (953) | 18.1 (264) | **16.5 (241)** |
| [81–max] | Female | 86.2 (631) | 10.5 (77) | **3.3 (24)** | 71.7 (525) | 20.1 (147) | **8.2 (60)** |
| | Female as male | 85.1 (622) | 9.4 (69) | **5.5 (40)** | 69.4 (507) | 17.1 (125) | **13.5 (99)** |

**Table S3. Comparison of age groups regarding AKI stages and sex. Based on common AKI cases for males and males as young.**

| Age group | | First AKI stage during hospitalization, %, (n) | | | Maximum AKI stage during hospitalization, %, (n) | | |
|---|---|---|---|---|---|---|---|
| | | AKIN1 | AKIN2 | AKIN3 | AKIN1 | AKIN2 | AKIN3 |
| [18–41) | Male | 74.2 (187) | 17.9 (45) | **7.9 (20)** | 57.5 (145) | 23.8 (60) | **18.7 (47)** |
| | Males as young | 76.6 (193) | 15.1 (38) | **8.3 (21)** | 59.5 (150) | 20.2 (51) | **20.2 (51)** |
| [41–61) | Male | 79.2 (795) | 12.6 (127) | **8.2 (82)** | 56.1 (563) | 21.9 (220) | **22.0 (221)** |
| | Males as young | 83.7 (840) | 8.5 (85) | **7.9 (79)** | 62.1 (623) | 17.1 (172) | **20.8 (209)** |
| [61–81) | Male | 82.7 (1827) | 10.5 (232) | **6.8 (150)** | 64.1 (1417) | 18.4 (407) | **17.4 (385)** |
| | Males as young | 83.7 (1848) | 5.6 (123) | **10.8 (238)** | 66.8 (1476) | 10.7 (236) | **22.5 (497)** |
| [81–max] | Male | 87.0 (578) | 7.5 (50) | **5.4 (36)** | 70.0 (465) | 15.5 (103) | **14.5 (96)** |
| | Males as young | 82.1 (545) | 2.7 (18) | **15.2 (101)** | 64.5 (428) | 7.8 (52) | **27.7 (184)** |
